# Genomic characterization of a novel SARS-CoV-2 lineage from Rio de Janeiro, Brazil

**DOI:** 10.1101/2020.12.23.20248598

**Authors:** Carolina M Voloch, Ronaldo da Silva F, Luiz G P de Almeida, Cynthia C Cardoso, Otavio J. Brustolini, Alexandra L Gerber, Ana Paula de C Guimarães, Diana Mariani, Raissa Mirella da Costa, Orlando C. Ferreira, Covid19-UFRJ Workgroup, LNCC-Workgroup, Adriana Cony Cavalcanti, Thiago Silva Frauches, Claudia Maria Braga de Mello, Rafael Mello Galliez, Débora Souza Faffe, Terezinha M P P Castiñeiras, Amilcar Tanuri, Ana Tereza R de Vasconcelos

## Abstract

In this study, we report the sequencing of 180 new viral genomes obtained from different municipalities of the state of Rio de Janeiro from April to December 2020. We identified a novel lineage of SARS-CoV-2, originated from B.1.1.28, distinguished by five single-nucleotide variants (SNVs): C100U, C28253U, G28628U, G28975U, and C29754U. The SNV G23012A (E484K), in the receptor-binding domain of Spike protein, was widely spread across the samples. This mutation was previously associated with escape from neutralizing antibodies against SARS-CoV-2. This novel lineage emerged in late July being first detected by us in late October and still mainly restricted to the capital of the state. However, as observed for other strains it can be rapidly spread in the state. The significant increase in the frequency of this lineage raises concerns about public health management and continuous need for genomic surveillance during the second wave of infections.

**Article Summary Line:** We identified a novel circulating lineage of SARS-CoV-2 in the state of Rio de Janeiro Brazil originated from B.1.1.28 lineage.

## Introduction

The first cases of the disease caused by the new coronavirus SARS-CoV-2 (COVID-19) were reported in Wuhan province, in China, in December 2019. A few months later, in March 2020, the virus had already spread worldwide, in a pandemic that has surpassed 76 million cases and 1,6 million deaths (https://coronavirus.jhu.edu/map.html) in one year.

Since a vaccine is still unavailable for most people, controlling viral spread can be quite challenging, especially in countries that have not implemented broad testing and isolation of active cases. In this scenario, transmission suppression depends mainly on the continuous population adherence to non-pharmacological strategies such as social distancing. In mid-December, when this manuscript was prepared, European countries were already facing a second wave of SARS-CoV-2 transmission. At the same time, Brazil was the country with the third highest absolute number of COVID-19 cases and the second in lethality, with more than 7,200,000 cases and 186,000 deaths (https://coronavirus.jhu.edu/map.html).

Rio de Janeiro is the third Brazilian State in total inhabitants. According to data from December 14^th^, although Rio de Janeiro was also the second in the absolute number of COVID-19 cases, it was the first in cumulative COVID-19 deaths in Brazil – 138 per 100,000 habitants. State’s capital accounts for the highest cumulative number of deaths in the State – 207 per 100,000 (https://susanalitico.saude.gov.br/). The active transmission sustained since March and the high number of cases offer a suitable scenario for genomic surveillance. Such investigation is crucial to define routes of viral spread, the outcome of new lineages, and possible variations in transmissibility/infectivity or attenuation of SARS-CoV-2.

We have previously demonstrated the early spread of SARS-CoV-2 in Brazil (1). Here, we report the evolution of the epidemic in Rio de Janeiro since the early beginning, in April, until shortly before the new increase in COVID-19 incidence in December. Based on the analysis of complete viral genome sequences obtained from 180 patients from different municipalities, our data show a rise in B.1.1.28 lineage frequency, from where a new variant lineage with five exclusive mutations has emerged and rapidly spread.

## Methods

### Sample collection

A total of 180 participants, 82 males and 98 females, from 19 municipalities located at Rio de Janeiro State, in Brazilian Southeast region, were enrolled in this cross-sectional study. The study was conducted in partnership with the State Health Department of Rio de Janeiro. Most patients are residents of the capital, Rio Janeiro (60,6%). Subjects were selected from April to November, during CDC epidemiological weeks 15 to 45, from the cohorts of the Center for COVID-19 diagnosis from Federal University of Rio de Janeiro and Central Laboratory Noel Nutels, both in Rio de Janeiro and from the Molecular Diagnostic Laboratory Doutor Francisco Rimolo Neto, in Maricá. Age distribution was highly variable, with a median of 39 years old (range 16 to 87) (**Appendix 1 Table 1**). Nasopharyngeal swabs were obtained from each participant and COVID-19 diagnosis was performed by RT-PCR using CDC protocol, using primers and probes for N1 and N2 targets. The present study was approved by the National Committee of Research Ethics and by the Ethics Committee from Hospital Universitário Clementino Fraga Filho (protocol numbers: 30161620.0.1001.5257 and 34025020.0.0000.5257).

### Next-generation sequencing and bioinformatics analysis

cDNA synthesis and viral whole-genome amplification were carried out following the Artic Network protocol (https://artic.network/ncov-2019). Amplicon libraries were prepared either using the TruSeq DNA Nano kit or Nextera DNA Flex kit (Illumina, USA). Sequencing was performed in a MiSeq System using MiSeq Reagent Kit v3 (Illumina, USA). Quality assessment of sequencing data was performed using FastQC (v0.11.4). Next, we filtered low-quality reads (<25) with trimmomatic v0.39 (2). The sequences were mapped to the reference genome NC_045512.2 using the BWA 0.7.17 software. We then generated the consensus genome sequence using bcftools v1.10.2 and bedtools v2.29.2 packages (3–5). Single-nucleotide variants (SNVs) were detected using the GATK v4.1.7.0 (6). We used the adegenet package in R to perform clustering and discriminant analysis of principal components (DAPC) (7,8).

### Phylogenetic and phylogeographic Analysis

Brazilian dataset contains the 180 new consensus genomes and 116 other Brazilian high-coverage genomes available in GISAID (https://www.gisaid.org/) with collection dates ranging from May to November (Data downloaded from GSAID December 03, 2020). We appended this Brazilian dataset to a global dataset including 1081 genomes sampled from GISAID to include one genome per country per week (based on sample collection day), from May to November 2020 (**Appendix 2**).

To obtain a Maximum Likelihood (ML) phylogeny, we ran IQTree 2.0.3 (9) under a GTR model of nucleotide substitution (10) with empirical base frequencies and invariant sites. This model configuration was selected using IQ-Tree’s ModelFinder tool. We screened our datasets for recombination using the Phi-test approach in SplitsTree (11,12) and found no recombination evidence. We identified three Clades, I, II, and III, in the ML tree as monophyletic clusters that encompass the great majority of Rio de Janeiro genomes with at least three or more sequences from Rio de Janeiro clustered within the same clade.

Then, ML phylogeny was estimated for each of the three clades. We assessed the temporal signal of each Clade dataset using TempEst 1.5.3 (13) and removed the outlier sequences. Root-to-tip linear regression coefficient of genetic divergence against sampling dates 0.51, 0.71 and 0.88, for Clade I, II and III, respectively (Figure S1). The time-calibrated phylogenies were estimated under a strict molecular clock in BEAST v.1.10.4 (14) using the GTR+I (10) nucleotide substitution model for Clades I and II, and HKY nucleotide substitution model (15) for Clade III. We assumed an exponential growth tree prior and specified a normal prior for the clock rate with mean = 8×10-4 and stdev = 0,1×10-5. We also used a discrete diffusion model (16) to infer the state of each internal node of the phylogeny, considering the location as a discrete trait with only three possible states: “RJ”, “Brazil”, and “Others”. The convergence of the MCMC chains, which were run at least for 20 million generations and sampled every 1000th step, was inspected using Tracer v.1.7.1 (17). Maximum clade credibility (MCC) summary trees were generated using TreeAnnotator v.1.10.4 (14).

We used a relaxed random walk diffusion model (18) available in BEAST 1.10 (14) with a Cauchy distribution to estimate trees whose internal nodes are associated with geographic coordinates. We ran an independent analysis for each Rio de Janeiro Clade identified previously, excluding all genomes for whose geographical coordinates were not known. We attributed each genome’s latitude and longitude to a point randomly sampled within the patient’s municipality of residence (Appendix 1 Table 1). MCMC chains were run for 10 million generations and logged every 1000th step, with convergence assessed using Tracer v1.7.1 (17). We used the spread3 package (19) to extract and map spatiotemporal information embedded in posterior trees.

## Results

On average, the viral genome assembly reached 99% of the sites coveraged, with a depth mean greater than 3500x (**Appendix 1 Table 2**). We found a total of 731 single-nucleotide variants (SNVs) across the 180 samples, of which 50.3% were missense, 44.5% synonymous, 5.1% intergenic and 0.1% nonsense (**Appendix 1 Table 3**). Most of the variants were identified exclusively in one sample with an allele frequency lower than 1%. The SNVs with allele frequencies greater than 60% corresponded to the mutational signatures of viral lineages derived from B.1.1 (**Figure 1A**). We also found ten variants with intermediate frequencies between 20-35% of which five occurred together in many samples.

**Figure 1.**
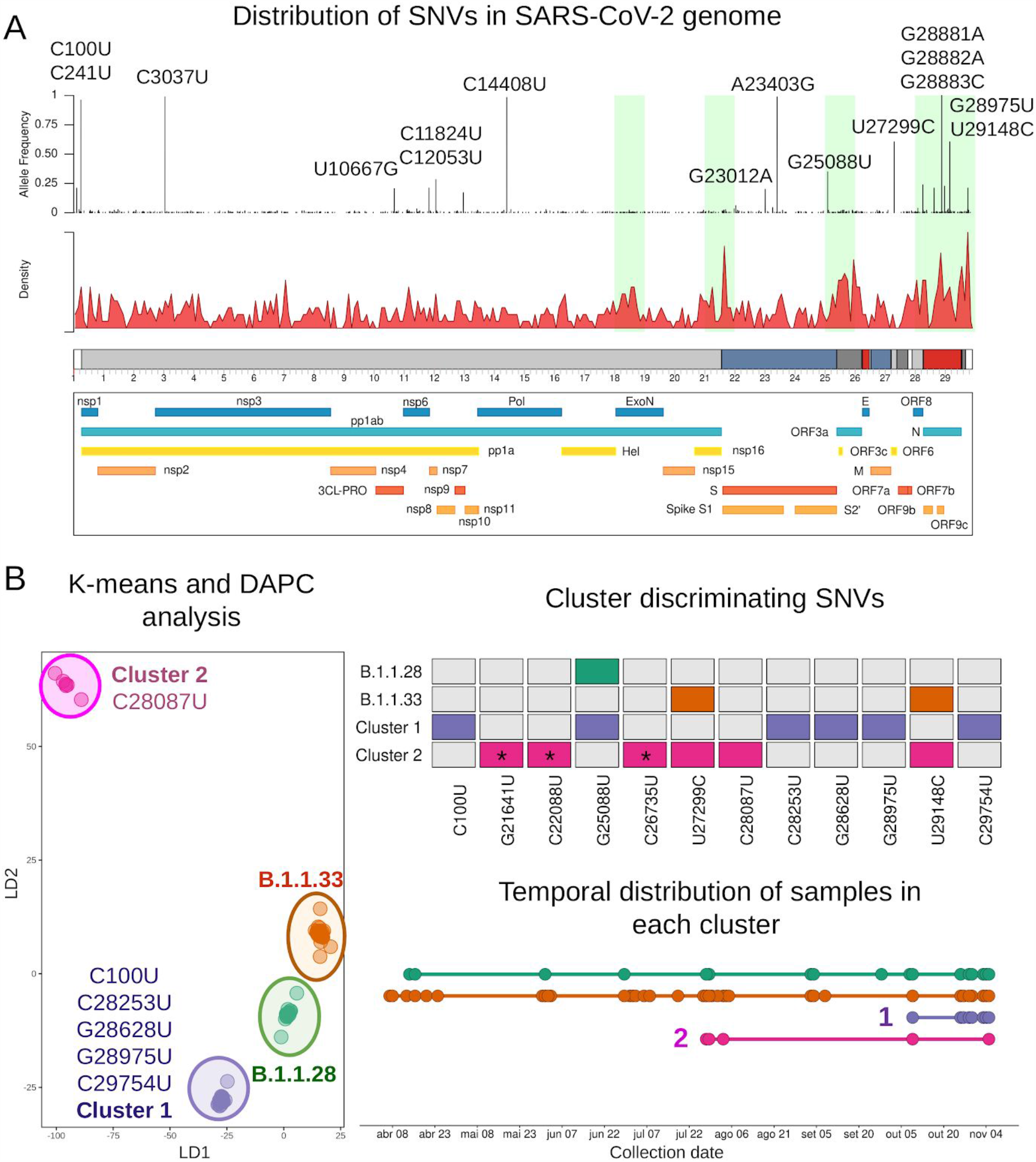
Genetic characterization and population structure analysis of SARS-CoV-2 genomes in Rio de Janeiro. A) Distribution of single-nucleotide variants across the viral genome. Each vertical line represents the variant frequency across the 180 samples targeting the protein products of the virus. Regions highlighted in green represent the top five kilobases with the highest number of mutations. In red, we showed the mutation density throughout the genome. B) K-means and discriminant analysis of principal components (DAPC) showing the four clusters detected by our analysis. Each cluster was characterized by at least one discriminant SNV. B1.1.28 and Cluster 1 shared one variant whereas B.1.1.33 and Cluster 2 had two SNVs in common. We also showed the temporal distribution of the samples from each group.

Per kilobase genetic screening revealed the highest mutation ratio in the 3’ end of the genome, mainly targeting ORF3a and N proteins (**Figure 1A**; **Appendix 1 Table 4**). These regions were also identified using a per-protein screening, in addition to ORF9c and other proteins (**Appendix 1 Table 5**). The high number of mutations found in ORF3a was due to the accumulation of non-synonymous variations throughout the genome (**Figure S2**). This pattern was found exclusively in ORF3a and S proteins, whereas a balanced acquisition of novel synonymous and non-synonymous SNVs was observed in N and ORF9c (**Figure S2**). Similar results were obtained in other structural and non-structural proteins of the SARS-CoV-2 genome (**Figure S2**).

B.1.1.33 (58%) and B.1.1.28 (32%) represented the overwhelming majority of viral lineages spread in Rio de Janeiro (**Appendix 1 Table 1**). Nevertheless, the co-occurrence of intermediate-frequency SNVs may suggest the emergence of novel variant lineages during the epidemic. We then applied a K-means algorithm combined with a discriminant analysis of principal components (DAPC) to ascertain the dynamic of population structure during the course of the epidemic in Rio de Janeiro. By retaining the first 100 principal components, we achieved approximately 90% of genetic variability in our samples (**Figure S3A**). Bayesian information criterion (BIC) values showed an optimal clusterization of four different groups: B.1.1.28 and B.1.1.33 lineages and two additional clusters, named Cluster 1 and Cluster 2 (**Figure 1B; Figure S3B**). Thirty-two out of the 180 samples were assigned to the B.1.1.28 cluster characterized by the missense variant G25088U (V1176F) in S protein, also shared among all samples from Cluster 1 (**Figure 1C**). The greatest number of samples (n=103) was observed in cluster B.1.1.33, represented by two missense variants U27299C (I33T) and U29148C (I292T) in ORF6 and N proteins, respectively.

Cluster 1 was composed of 38 samples, predominantly from B.1.1.28 lineage, detected after early October (**Figure 1D**). This cluster was characterized by the co-occurrence of five exclusive variants. Two variants mapped the 5’ and 3’ end (C100U and C29754U) respectively, a synonymous C28253U (F120F) in ORF8, and two missense G28628U (A119S) and G28975U (M234I) in N protein. In addition to these five variants, we also found the substitution G23012A (E484K) in S protein across 36 out of the 38 samples in this group. In cluster 2, one exclusive missense variant C28087U (A65V) in ORF8 was found across all seven samples. We also observed three SNVs G21641U (A27S), C22088U (L176F), and C26735U (Y71Y) targeting S and M in four out of the seven samples from this cluster. Samples from B.1.1.28 lineage and Cluster 1 were predominantly found in samples collected close to the state’s capital, whereas B.1.1.33 and Cluster 2 were also detected in other regions of the state (**Appendix 1 Table 1**).

Next, we tested the monophyletic status of Clusters 1 and 2 to identify whether they are possible emerging lineages. The phylogenetic reconstruction of a worldwide dataset with more than a thousand viruses confirmed that the 38 genomes from Cluster 1 formed a well supported monophyletic clade located inside a larger clade composed of B.1.1.28 strains (**Figure 2**). The monophyletic condition was not observed for the seven samples of Cluster 2, although they are all within clade B.1.1.33.

**Figure 2:**
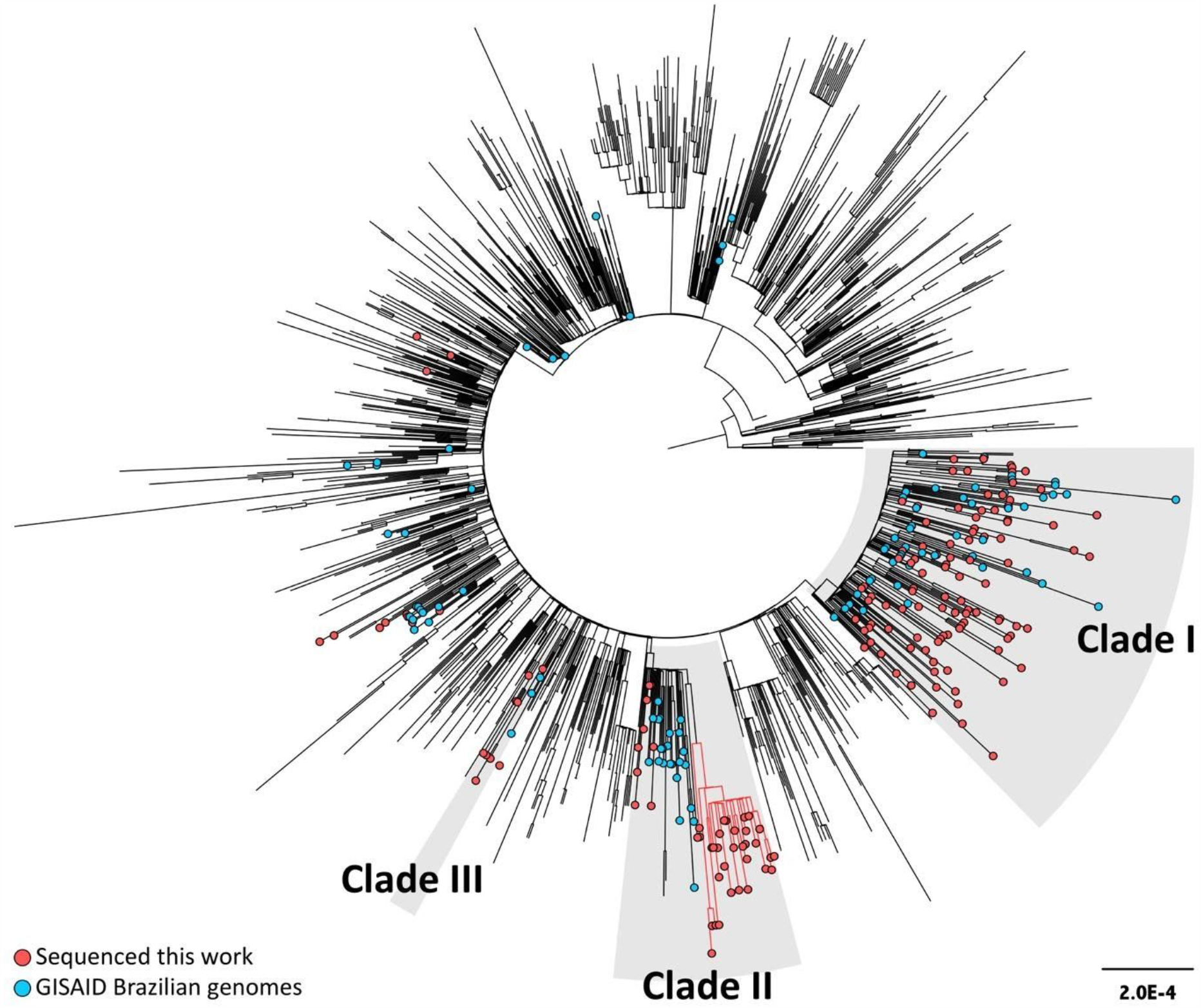
Maximum Likelihood tree of 1376 SARS-CoV-2 genomes including the 180 new Brazilian genomes generated in this work. We randomly sampled 1 genome/week/country from all available samples in GISAID with collection dates ranging from May to November. Brazilian sequences with collection dates from May to November were all added to the dataset. Tips are colored according to their origin. Genomes generated from this work are red, other Brazilian genomes are Blue, and genomes from other countries are not colored. Grey boxes represent the three clades where Brazilian viruses are concentrated. The emergent lineage identified in this work is highlighted with red branches.

We also observed that the vast majority (86.8%) of Brazilian genomes collected between May and November 2020 (257 out of 296) fall within three clades named hereafter Clade I, Clade II, and Clade III (Figure 2). Clade I is composed of B.1.1.33 strains, while Clade II and III are composed of B.1.1.28 viruses. Other lineages, like B.1 and B.1.1, were also sampled but had a minor contribution to establishing the epidemics in Brazil. None of those three lineages appear within the strains composing any of the three clades. Clade I encompasses 55% of the Brazilian genomes (165 out of 296), followed by Clade II with 26% and Clade III with 4%. Rio de Janeiro strains were also almost restricted to those clades (200 out of 220; 90.9%), with 63.2% of Rio de Janeiro strains within Clade I; 24.1% within Clade II; and 3.6% within Clade III. Further analyses were only performed with those three clades as they contain the vast majority of samples from Rio de Janeiro.

Emergence of Cluster 1 was dated around early July 2020 (Figure 3). Interestingly, the most recent sample from the new variant lineage dates from the late of October, almost four months after it’s estimated emergence. A clear pattern of clustering can be observed from Rio de Janeiro viruses, since they appear to be more related to each other than to the different localities. Even in the complete dataset analysis, it is possible to observe clades exclusively from Rio de Janeiro (Figure 2). We inferred the location of ancestral nodes with our discrete phylogeographic analysis and, as expected, the state inferred for most internal nodes in the three clades is Rio de Janeiro (Figure 3). Most of our samples are within clades whose ancestral location is Rio de Janeiro, and the clades with Rio de Janeiro ancestral state are almost exclusively composed of viruses from that location.

**Figure 3:**
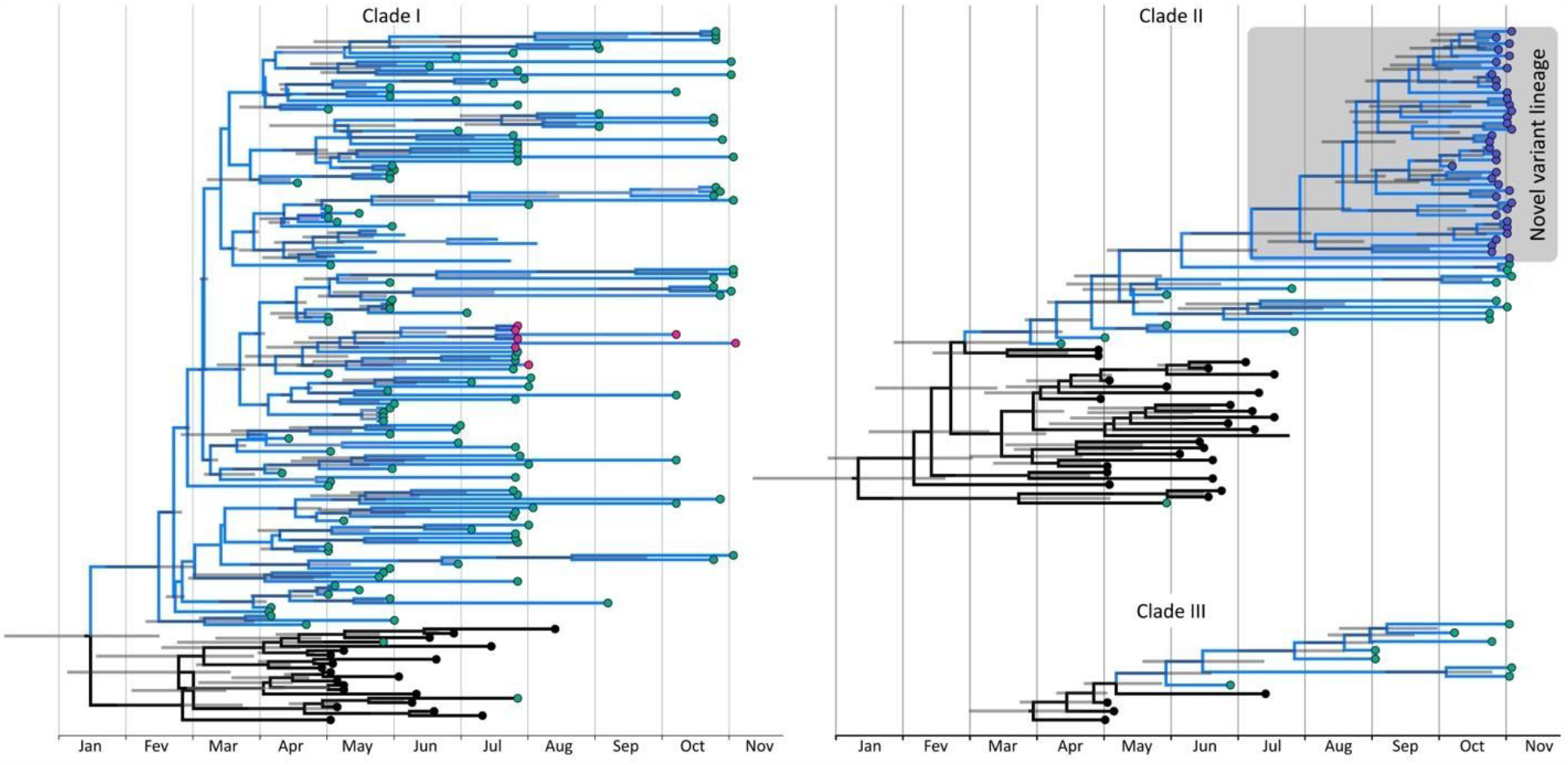
Time scaled trees of the tree Brazilian Clades of SARS-CoV-2. Circles in the tip nodes are colored according to location. Samples from Rio de Janeiro are shown in green circles, pink represents cluster 2 and purple the novel lineage identified in this work, that is also highlighted in a grey box. Substitutions that characterize the emergent lineage are shown in blue above the branch defining the lineage. Grey bars represent 95% hpd interval for the inferred ages. Rio de Janeiro clades identified by the discrete phylogeographic analysis are shown in blue.

We performed a continuous phylogeographic reconstruction to model the virus spread across the state (**Figure 4**). We estimated that the Rio de Janeiro city represented the primary source of virus strains circulating within the state. Our reconstructions also reveal the occurrence of few long-distance dispersal events from the capital to more remote areas of the state. We inferred few lineage movements towards the state’s capital, all from the Maricá city.

**Figure 4:**
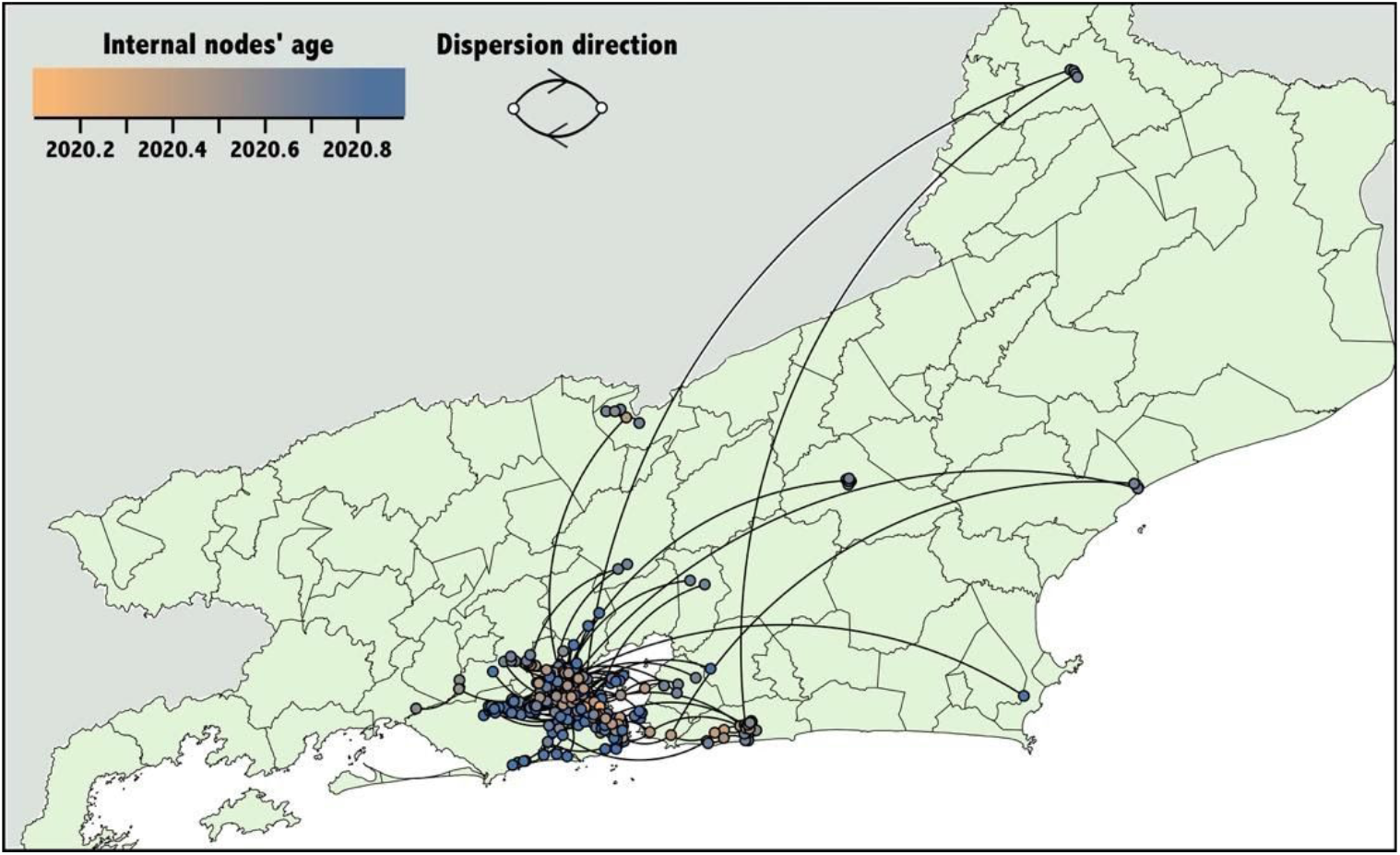
Phylogeographic inference of the Spread of SARS-CoV-2 in Rio de Janeiro state. Spatially-explicit reconstruction of Rio de Janeiro SARS-CoV-2 clusters identified by the discrete phylogeographic analysis in each of the tree Clades. Each dot represents a node of the MCC tree and are coloured according to their inferred time of occurrence. Curved lines represent branches of phylogeny and the directionality of movement as depicted in the figure legend.

## Discussion

In this study, we identified a novel circulating strain of SARS-CoV-2 originated from B.1.1.28 lineage and widely spread in Rio de Janeiro state. We estimate that the novel lineage emerged in early July, approximately four months before the detection of the first genomes from this lineage. We demonstrated that the COVID-19 epidemic in Rio de Janeiro was characterized by three main clades, of which two represented the vast majority of the circulating viruses, classified as B.1.1.28 and B.1.1.33 lineages. Interestingly, this division of three clades was also observed in Brazil during the beginning of the SARS-CoV-2 pandemic (1).

According to our results, Rio de Janeiro sequences were also mainly restricted to those three clades, and more often related to each other than to strains from other states or outside the country. This pattern suggests a limited contribution of foreign viruses to the composition of Rio de Janeiro lineages. However, these results could be biased due to the scarcity of Brazilian samples. Whether this aggregation pattern of Rio de Janeiro genomes with few inferred introduction events is due to the high communitarian transmission, or a consequence of low sampling, needs further investigation. Despite the low sampling ratio, our data indicate that the transmission chains circulating in Rio de Janeiro were established by a restricted number of infectious cases, a fact that was not observed in other reports where there is an important contribution of external introduction events and no predominant lineage was identified (20,21).

We noticed an alteration in the relative proportion of genomes sampled from each lineage in Brazil through time. Analyzing our data together with the genomes available in GISAID, we observed that B.1.1.28 samples are becoming more frequent in the country (Figure 5). In addition, the increase in the B.1.1.28 relative frequency is synchronous with the age inferred for the ancestral of the novel variant lineage. The emergence of this lineage can probably be associated with the elevation in the number of the COVID-19 cases (so called second wave) in the state. Together with the pre-existing circulating viral lineages in Rio de Janeiro, the second wave of infection may be characterized by the presence of this emergent lineage.

**Figure 5:**
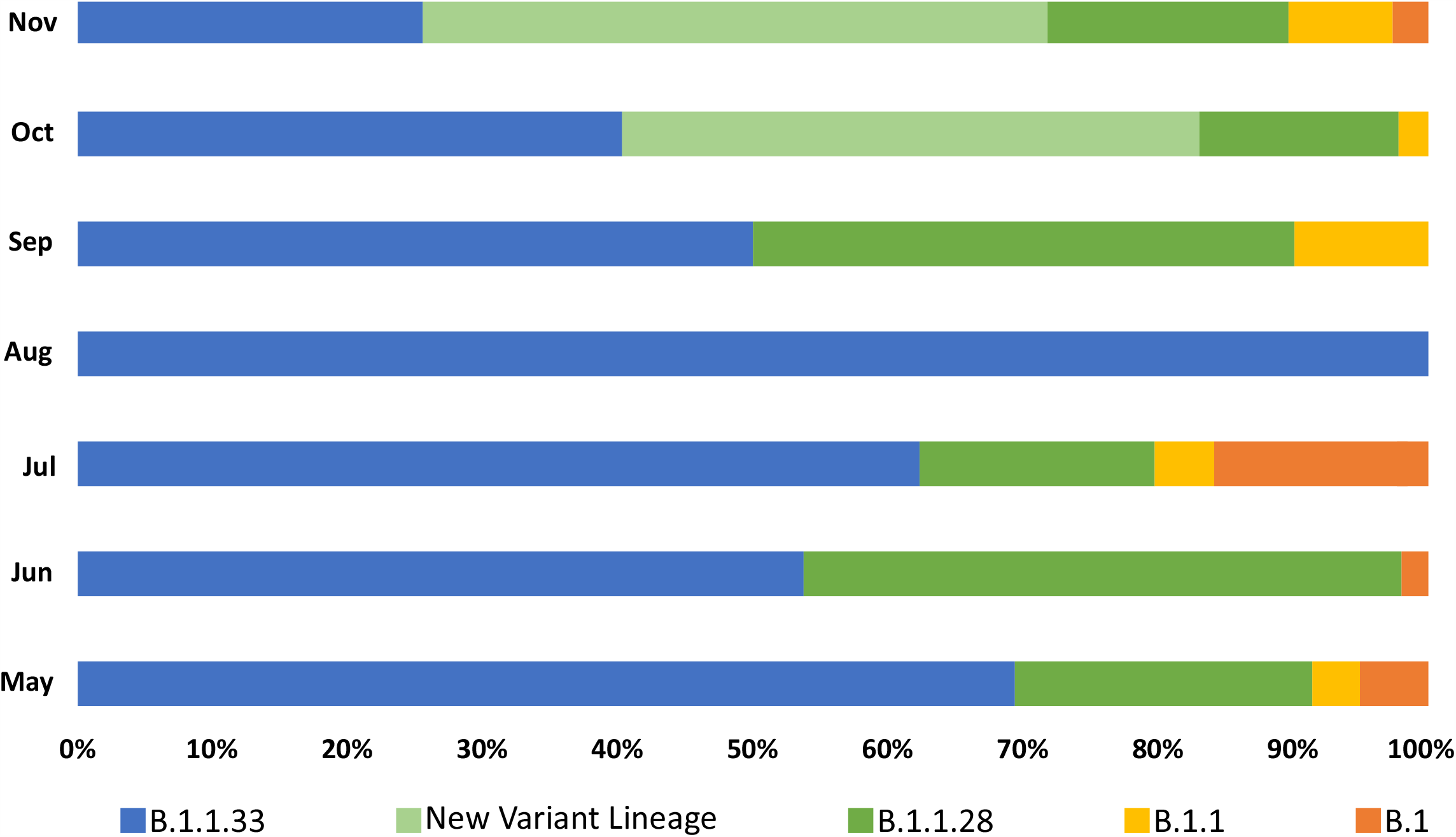
Evolution of the relative frequency of Brazilian SARS-CoV-2 lineages. A Brazilian dataset composed of the 180 genomes generated in this work plus all Brazilian genomes with collection dates ranging from May to November available in GISAID, was submitted to Pangolin classification. The relative frequency of each lineage in each month was plotted. Colors represent each lineage as described in the figure legend.

Many mutations arise frequently in the SARS-CoV-2 genome during the virus replication cycle and intra-host defense mechanisms (22). However, most of these variants are lost as a result of the natural selection process, which also lead to fixation of some variants in the population. The co-occurence of fixed mutations in the population will characterize the novel viral lineage.

Indeed, novel SARS-CoV-2 lineages are arising in several regions around the world (23). Recently, phylogenetic analysis revealed a novel emergent SARS-CoV-2 lineage in the United Kingdom mainly distinguished by the variant A23063T (N501Y) targeting the receptor-binding domain (RBD) in Spike protein. Interestingly, beside the five SNVs that characterized our novel lineage, we also identified the mutation G23012A (E484K) in the RBD region of Spike protein. This alteration was previously associated with the escape from neutralizing antibodies against SARS-CoV-2 (24,25). Nevertheless, further analyses are required to predict whether the changes in these novel lineages have an important effect on viral infectivity, host immune response or disease severity. Experimental analyses are also needed to understand the molecular and physiological implications of these variations.

Efforts to continuously monitor the virus evolution during the course of the pandemic are crucial to assess the dynamic of infection and population structure. Nonetheless, genomic surveillance is still being a challenge in Brazil given the low number of sequences generated during this period. Our study has provided a significant increase in the number of genomes available in public databases from Rio de Janeiro and consequently from Brazil (adding 180 new near complete genomes to the 314 already available in GISAID). Finally, as of 18 Dezember 2020, when we were writing this work, four sequences harboring the same five exclusive variants of the new variant lineage were already found in the United Kingdom and United States (GISAID accession numbers EPI_ISL_668335, EPI_ISL_677211, EPI_ISL_677212, EPI_ISL_683969). The collection dates were all more recent than ours (ranging from 2020-11-12 to 2020-11-14). Ultimately, this may indicate the introduction of this Brazilian lineage worldwide.

## Data availability

NGS data generated in our study is publicly available in SRA-NCBI (www.ncbi.nlm.nih.gov/sra), Bioproject accession PRJNA686081. Genome sequences are also deposited in Gisaid (www.gisaid.org) and the access identifiers are listed in Appendix 1 table 6.

## Acknowledgments

This work was developed in the frameworks of Corona-ômica-RJ (FAPERJ = E-26/210.179/2020) and Rede Corona-ômica BR MCTI/FINEP (FINEP = 01.20.0029.000462/20, CNPq = 404096/2020-4). The study was also supported by FAPERJ E-26/010.002434/2019 and E-26/210.178/2020 and Serrapilheira Institue, Brazil for A.T, and E-26/010.002278/2019 for C.M.V. A.T.R.V. is supported by Conselho Nacional de Desenvolvimento Científico e Tecnológico - CNPq (303170/2017-4) and FAPERJ (E-26/202.903/20). R.S.F.J is a recipient of a graduate fellowship from CNPq. C.C.C is supported by FAPERJ (E-26/202.791/2019).

## Workgroup Members

### Covid19-UFRJ Workgroup

Alexandre Otávio Chieppe, Mario Sergio Ribeiro, Silvia Cristina de Carvalho Cardoso, João de Farias Figueiredo, Leandro Magalhães de Souza, Veronica Guedes de Nogueira, Frederico Francisco Nascimento, Jose Paulo Motta Filho, Isabela de Carvalho Leitão, Anna Carla Pinto Castiñeiras, Romina Carvalho Ferreira, Bianca Ortiz da Silva, Helena D’Anunciação de Oliveira, Guilherme Sant’Anna de Lira, Helena Toledo Scheid, Laura Zalcberg Renault, Leticia Averbug Correa, Marcela dos Santos Durães, Raquel Fernandes Coelho, Victor Akira Ota, Victoria Cortes Bastos, Roberto Medronho, Lucas Dalsenter Romano da Silva, Carlos Alberto de Senna Costa, Celso Pansera, Claudia dos Santos Rodrigues, Andreia Ribeiro Costa, Wilson Rodrigues de Souza Junior, Marcelo Costa Velho Mendes de Azevedo, Luciana Jesus Costa, Alice Laschuk Herlinger, Cassia Cristina Alves Gonçalves, Cíntia Policarpo, Érica Ramos dos Santos Nascimento, Fernanda Leitão dos Santos, Filipe Romero Rebello Moreira, Francine Bittencourt Schiffler, Jessica Maciel de Almeida, Juliana Tiemi Sato Fortuna, Lídia Theodoro Boullosa, Marcelo Calado de Paula Tôrres, Mirela D’arc Ferreira da Costa, Pedro Henrique Costa da Paz, Thais Felix Cordeiro da Cruz, Ricardo José Barbosa Salviano.

## LNCC-Workgroup

Luciane Prioli Ciapina, Rangel Celso Souza, Éllen dos Santos Correa, Bruno Zonovelli da Silva, Guilherme Cordenonsi da Fonseca, Vinícius Prata Klôh, Eduardo Wagner, Liliane Tavares de Faria Cavalcante.

## Disclaimers

Authors have disclosed any conflicts of interest related to this article.

## Author Bio

Carolina M Voloch is associate professor at the Genetics department of Federal University of Rio de Janeiro. Research focuses on use of bioinformatics tools to study the molecular evolution, phylogenetics and genomics of viruses.

Ronaldo da Silva Francisco Junior is member of the Laboratório de Bioinformática (LABINFO/LNCC) and PhD candidate in Genetics at the Federal University of Rio de Janeiro. The most recent background is in bioinformatics, genetic variant analysis and clinical genomics, focusing on variant calling, whole exome sequencing and RNA-Seq.

## Figure

**Figure S1.**
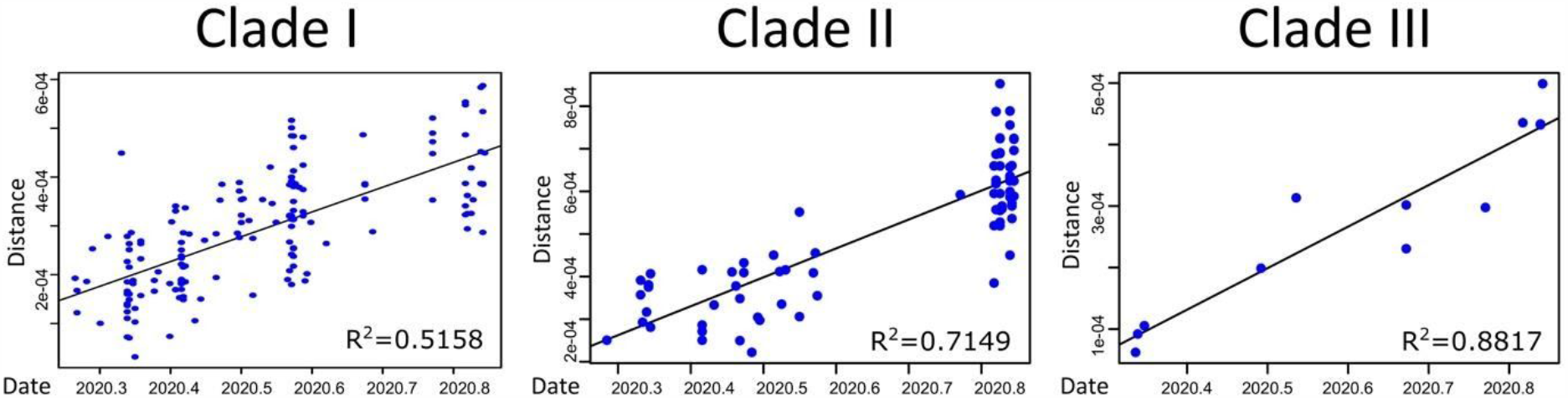
Root-to-tip regression of genetic divergence against dates of sample collection for Clade I, II and III datasets. The coefficients of the linear regression are also shown for each case.

**Figure S2.**
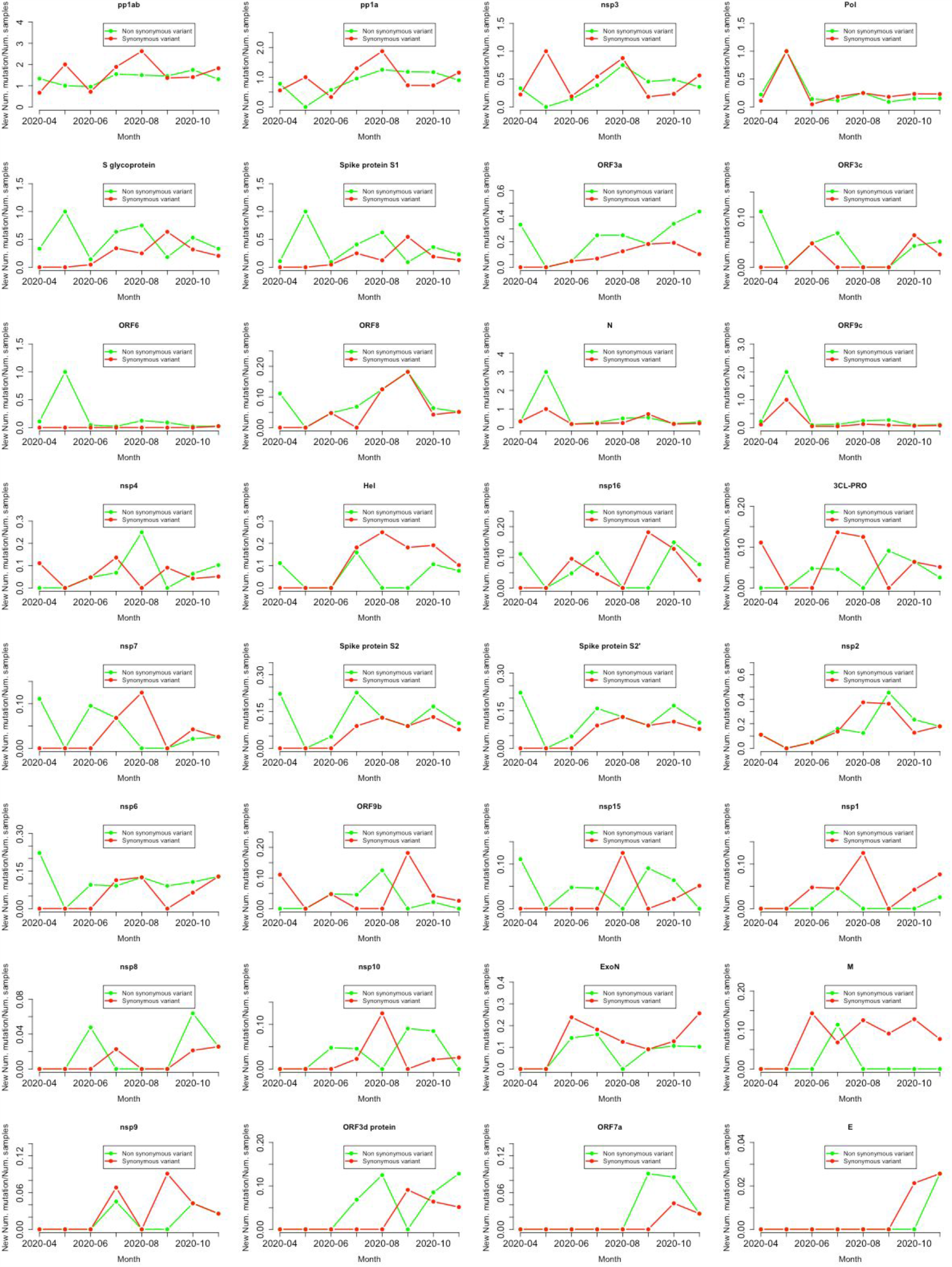
Acquisition of synonymous and non-synonymous variants in each protein product of SARS-CoV-2 genome over time. Monthly acquisition of different variants in each protein of the virus over time. The x-axis represents the interval of months from April to November and y-axis the number of novel variants identified normalized by the number of samples sequenced in each month.

**Figure S3.**
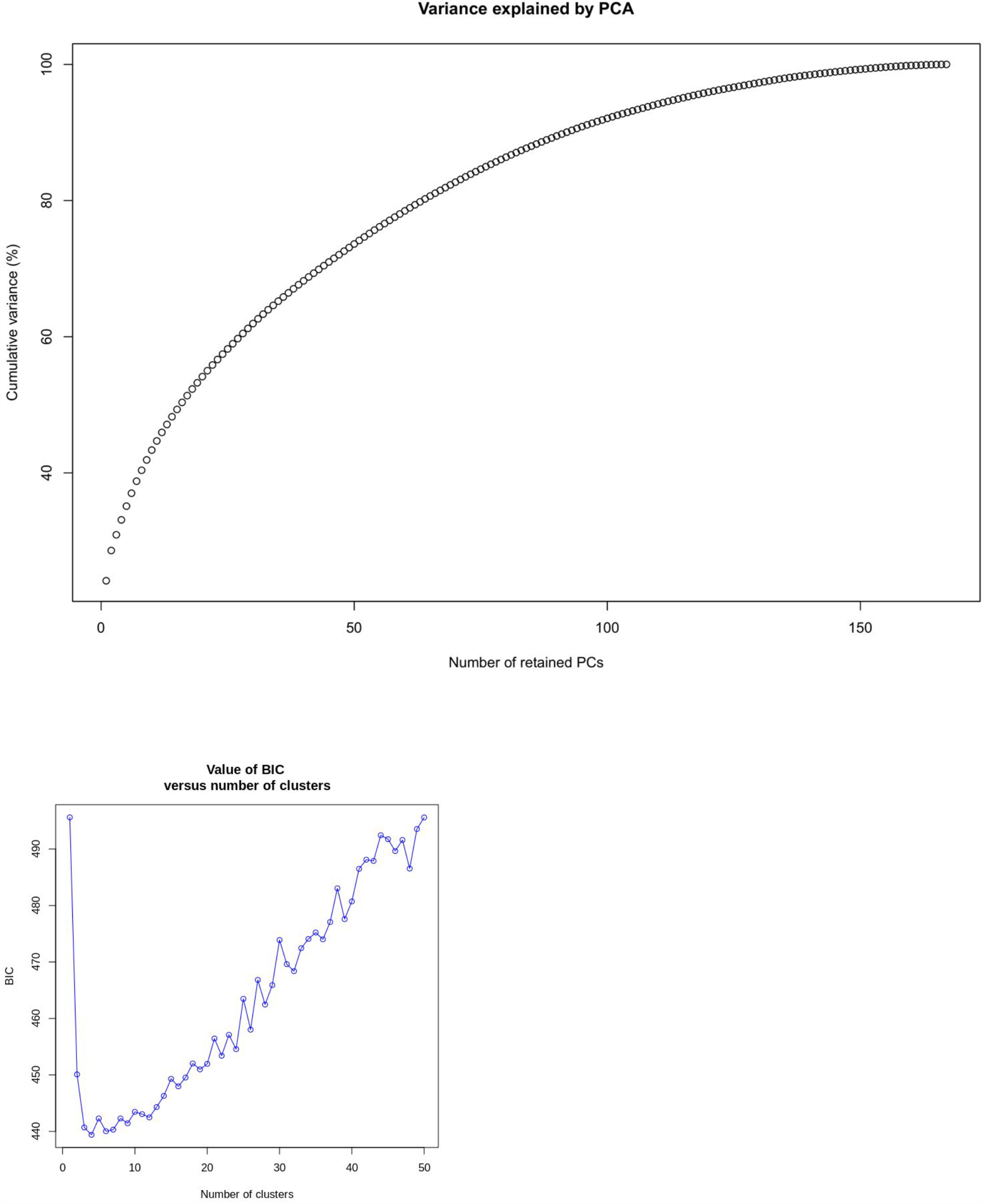
Genetic variability and population structure inference. A) Principal component analysis (PCA) of the genetic diversity found in the 180 samples. B) Bayesian information criterion (BIC) for each possible number of cluster.

## Tables

**Appendix 1 Table 1**: Epidemiology and sample information.

**Appendix 1 Table 2**: Quality control and of sequencing metrics.

**Appendix 1 Table 3**: Genomic variants identified across the SARS-CoV-2 genome in the 180 samples sequenced.

**Appendix 1 Table 4**: Per-kilobase mutation ratio analysis.

**Appendix 1 Table 5**: Per-protein mutation ratio analysis.

**Appendix 1 Table 6**: Identifiers to access data in NCBI/SRA and GISAID

**Appendix 2 :** GISAID acknowledgement table

## References

1. Candido DS, Claro IM, de Jesus JG, Souza WM, Moreira FRR, Dellicour S, et al. Evolution and epidemic spread of SARS-CoV-2 in Brazil. Science. 2020 Sep 4;369(6508):1255–60.

2. Bolger AM, Lohse M, Usadel B. Trimmomatic: a flexible trimmer for Illumina sequence data [Internet]. Vol. 30, Bioinformatics. 2014. p. 2114–20. Available from: http://dx.doi.org/10.1093/bioinformatics/btu170

3. Li H. A statistical framework for SNP calling, mutation discovery, association mapping and population genetical parameter estimation from sequencing data. Bioinformatics. 2011 Nov 1;27(21):2987–93.

4. Li H. Improving SNP discovery by base alignment quality. Bioinformatics. 2011 Apr 15;27(8):1157–8.

5. Quinlan AR, Hall IM. BEDTools: a flexible suite of utilities for comparing genomic features. Bioinformatics. 2010 Mar 15;26(6):841–2.

6. DePristo MA, Banks E, Poplin R, Garimella KV, Maguire JR, Hartl C, et al. A framework for variation discovery and genotyping using next-generation DNA sequencing data. Nat Genet. 2011 May;43(5):491–8.

7. Jombart T. adegenet: a R package for the multivariate analysis of genetic markers [Internet]. Vol. 24, Bioinformatics. 2008. p. 1403–5. Available from: http://dx.doi.org/10.1093/bioinformatics/btn129

8. Jombart T, Ahmed I. adegenet 1.3-1: new tools for the analysis of genome-wide SNP data. Bioinformatics. 2011 Nov 1;27(21):3070–1.

9. Minh BQ, Schmidt HA, Chernomor O, Schrempf D, Woodhams MD, von Haeseler A, et al. IQ-TREE 2: New Models and Efficient Methods for Phylogenetic Inference in the Genomic Era. Mol Biol Evol. 2020 May 1;37(5):1530–4.

10. Tavaré S. Some probabilistic and statistical problems in the analysis of DNA sequences. Lectures on Mathematics in the Life Sciences. 1986;17:57–86.

11. Bruen TC, Philippe H, Bryant D. A simple and robust statistical test for detecting the presence of recombination. Genetics. 2006 Apr;172(4):2665–81.

12. Huson DH, Bryant D. Application of Phylogenetic Networks in Evolutionary Studies [Internet]. Vol. 23, Molecular Biology and Evolution. 2006. p. 254–67. Available from: http://dx.doi.org/10.1093/molbev/msj030

13. Rambaut A, Lam TT, Max Carvalho L, Pybus OG. Exploring the temporal structure of heterochronous sequences using TempEst (formerly Path-O-Gen). Virus Evol. 2016 Jan;2(1):vew007.

14. Suchard MA, Lemey P, Baele G, Ayres DL, Drummond AJ, Rambaut A. Bayesian phylogenetic and phylodynamic data integration using BEAST 1.10. Virus Evol. 2018 Jan;4(1):vey016.

15. Hasegawa M, Kishino H, Yano T. Dating of the human-ape splitting by a molecular clock of mitochondrial DNA. J Mol Evol. 1985;22(2):160–74.

16. Lemey P, Rambaut A, Drummond AJ, Suchard MA. Bayesian phylogeography finds its roots. PLoS Comput Biol. 2009 Sep;5(9):e1000520.

17. Rambaut A, Drummond AJ, Xie D, Baele G, Suchard MA. Posterior Summarization in Bayesian Phylogenetics Using Tracer 1.7. Syst Biol. 2018 Sep 1;67(5):901–4.

18. Lemey P, Rambaut A, Welch JJ, Suchard MA. Phylogeography takes a relaxed random walk in continuous space and time. Mol Biol Evol. 2010 Aug;27(8):1877–85.

19. Bielejec F, Baele G, Vrancken B, Suchard MA, Rambaut A, Lemey P. SpreaD3: Interactive Visualization of Spatiotemporal History and Trait Evolutionary Processes. Mol Biol Evol. 2016 Apr 23;33(8):2167–9.

20. Dellicour S, Durkin K, Hong SL, Vanmechelen B, Martí-Carreras J, Gill MS, et al. A Phylodynamic Workflow to Rapidly Gain Insights into the Dispersal History and Dynamics of SARS-CoV-2 Lineages. Mol Biol Evol [Internet]. 2020 Nov 3; Available from: http://dx.doi.org/10.1093/molbev/msaa284

21. Deng X, Gu W, Federman S, du Plessis L, Pybus OG, Faria N, et al. A Genomic Survey of SARS-CoV-2 Reveals Multiple Introductions into Northern California without a Predominant Lineage. medRxiv [Internet]. 2020 Mar 30; Available from: http://dx.doi.org/10.1101/2020.03.27.20044925

22. Voloch CM, da Silva F R, de Almeida LGP, Brustolini OJ, Cardoso CC, Gerber AL, et al. Intra-host evolution during SARS-CoV-2 persistent infection. medRxiv. 2020 Nov 16;2020.11.13.20231217.

23. Hodcroft EB, Zuber M, Nadeau S, Crawford KHD, Bloom JD, Veesler D, et al. Emergence and spread of a SARS-CoV-2 variant through Europe in the summer of 2020. medRxiv. 2020 Nov 27;2020.10.25.20219063.

24. Greaney AJ, Starr TN, Gilchuk P, Zost SJ, Binshtein E, Loes AN, et al. Complete Mapping of Mutations to the SARS-CoV-2 Spike Receptor-Binding Domain that Escape Antibody Recognition. Cell Host Microbe [Internet]. 2020 Nov 19; Available from: http://dx.doi.org/10.1016/j.chom.2020.11.007

25. Weisblum Y, Schmidt F, Zhang F, DaSilva J, Poston D, Lorenzi JCC, et al. Escape from neutralizing antibodies by SARS-CoV-2 spike protein variants [Internet]. Cold Spring Harbor Laboratory. 2020 [cited 2020 Dec 20]. p. 2020.07.21.214759. Available from: https://www.biorxiv.org/content/10.1101/2020.07.21.214759v1.abstract

